# Strategies to Estimate Prevalence of SARS-CoV-2 Antibodies in a Texas Vulnerable Population: Results from Phase I of the Texas Coronavirus Antibody REsponse Survey (TX CARES)

**DOI:** 10.1101/2021.08.04.21261613

**Authors:** Melissa Valerio-Shewmaker, Stacia DeSantis, Michael Swartz, Ashraf Yaseen, Michael O. Gonzalez, Harold W. Kohl, Steven H. Kelder, Sarah E. Messiah, Kimberly A. Aguillard, Camille Breaux, Leqing Wu, Jennifer Shuford, Stephen Pont, David Lakey, Eric Boerwinkle

**Affiliations:** University of Texas Health Science Center, School of Public Health, Brownsville Campus; University of Texas Health Science Center, School of Public Health, Austin Campus, Austin, TX, USA; University of Texas Health Science Center, School of Public Health, Dallas Campus, Dallas, TX, USA; Texas Department of State Health Services, Austin, TX USA; University of Texas System, Population Health, Austin, Texas, USA

## Abstract

**Introduction:** Severe acute respiratory syndrome coronavirus 2 (SARS-CoV-2) infection and immunity remains uncertain in populations. The state of Texas ranks 2^nd^ in infection with over 2.71 million cases and has seen a disproportionate rate of death across the state. The TX CARES project was funded by the state of Texas to estimate the prevalence of SARS-CoV-2 antibody status in children and adults.

**Materials and Methods:** The TX CARES (Texas Coronavirus Antibody Response Survey) is an ongoing prospective population-based convenience sample from the Texas general population that commenced in October 2020. Volunteer participants are recruited across the state to participate in a 3-time point data collection TX CARES to assess antibody response over time. We use the Roche Elecsys® Anti-SARS-CoV-2 Immunoassay to determine SARS-CoV-2 antibody status.

**Results:** The crude antibody positivity prevalence in Phase I was 26.1% (80/307). The fully adjusted seroprevalence of the sample was 31.5%. Specifically, 41.1% of males and 21.9% of females were seropositive. For age categories, 33.5% of those 18-34; 24.4% of those 35-44; 33.2% of those 45-54; and 32.8% of those 55+ were seropositive. In this sample,42.2% (89/211) of those negative for the antibody test reported having had a COVID-19 test.

**Conclusions:** In this survey we enrolled and analyzed data for 319 participants, demonstrating a high survey and antibody test completion rate, and ability to implement a questionnaire and SARS-CoV-2 antibody testing within FQHC clinical settings. We were also able to determine our capability to estimate the cross-sectional seroprevalence within Texas’s FQHC clinical settings. The crude positivity prevalence for SARS-CoV-2 antibodies in this sample was 26.1% indicating potentially high exposure to COVID-19 for FQHC clinic employees and patients. These methods are being used to guide the completion of a large longitudinal survey in the state of Texas with implications for practice and population health.

## Introduction

Since January 2020, the Centers for Disease Control (CDC) has recommended county and state level reporting of all laboratory-confirmed cases of SARS-CoV-2 infection^1^. However, reported cases likely represent only a fraction of SARS-CoV-2 infections across the United States, as a still unknown proportion of cases are mild or asymptomatic ^2-5^, especially in young adults or children ^5-10^. Other challenges for SARS-CoV-2 surveillance include under-reported cases due to local health department capacity, delays in recording of testing and various methods of test reporting.^2-4, 11^ To obtain a more accurate representation of infection, many states and countries have turned to estimating SARS-CoV-2 seroprevalence from blood antibody assays allowing for an estimate of the prevalence of the human antibody response.^11-14^

Published data from the COVID-19 first and second wave indicate infections rates vary widely among different populations and geographic regions within a state^11^. Highly exposed populations include front line essential workers such as health care workers, teachers and educational staff, and those working in service, business, and retail, including grocery stores.^4^ Furthermore, ethnic minorities are at higher risk of contracting COVID-19^7,8^ as are vulnerable populations such as those without health insurance, people experiencing homelessness, or those with pre-existing conditions such as type 2 diabetes, hypertension and asthma. ^15-17^ Black and Latino communities have been especially hard hit by COVID-19 ^18-20^; for example, in a NYSDOH convenience sample of 15,000 New Yorkers *employed* at 99 grocery stores across 26 counties representing 87.3% of the state’s population found an adjusted seroprevalence ranging from 8.1% in non-Hispanic whites to 29.2% in Latinos, with an overall seroprevalence in New York City of 22.7%, versus a state-wide prevalence of only 8.9%. Other large seroprevalence studies are being conducted in California, Colorado, Georgia and Ohio ^11, 18, 20^.

Texas is the second largest state in the country and has a diverse population over 28,250,000 and a majority minority, with approximately 40% of residents identifying as Hispanic ethnicity, 12% as Black non-Hispanic and 7% other ethnicity. More than 34% of Texans live below 200% of federal poverty level (FPL). Texas is geographically diverse with approximately 85% of residents living in urban centers with vast rural areas requiring over one hour of travel to regional hospital systems^21^. Several areas of Texas have seen a high incidence of confirmed coronavirus disease (COVID-19) cases across two surges (July and December), including Dallas, Harris, Nueces, Cameron and Hidalgo counties. Furthermore, the prevalence of confirmed COVID-19 varies significantly across the state and by employment industry. For example, higher proportions of confirmed tests have been observed in underserved urban areas such as Dallas and Houston ^22-23^ and in areas with a high prevalence of vulnerable or Latino populations, such as San Antonio and McAllen, and in areas with multi-generational households, where viral transmission may be increased due to higher household density and with varied age groups within one household. We also see disparate burden of infection in rural areas with immigration detention centers (Willacy Co.) and meatpacking plants in the Texas Panhandle region^23^.

To ascertain estimate exposure to SARS-CoV-2 in the state of Texas, and to obtain an understanding of exposure across Texas, the Texas Coronavirus Antibody Research survey (Texas CARES) was designed as a longitudinal antibody surveillance study using a convenience sample approach from among highly exposed populations. Phase I of Texas CARES was designed to identify the feasibility of partnering and reaching vulnerable patients at Federally Qualified Health Centers (FQHCs) and to estimate seroprevalence of the 319 participants in this phase. There are currently 73 FQHCs serving patients in Texas, operating more than 500 sites and two FQHC lookalikes which offer FQHC-like services. The FQHCs are located across 126 counties and serve over 400,000 Medicaid patients, 28% of all FQHC patients, with 1,426,019 million patients served annually and over 5,300,000 patient visits annually^24^. We report here our Phase I sub-study of seroprevalence in a sample of 319 adults enrolling at three FQHC sites in Texas.

## Methods

All study protocols were reviewed and approved by the University of Texas Health Science Center Houston Institutional Review Board prior to any data collection. The Texas CARES program is a partnership with Texas Department of State Health Services and the University of Texas System with a statewide laboratory partner, Clinical Pathology Laboratories (CPL). The Texas Association of Community Health Centers (TACHC) partnered with us to introduce the program to FQHC sites. In total 40 or more FQHCs will be enrolled in the program over time.

### Study Population

The Phase I sub study of 319 participants presenting at or working at three FQHCs was performed as part of the larger Texas CARES study. The larger study aims to enroll participants from four populations across the state of Texas; pediatric school children 5-17 years of age, FQHC or community clinic patients, kindergarten to -12^th^ grade educators and allied staff and Texas workforce employees who will be tested for SARS-CoV-2 antibodies at three points over a 6-12 month period. The Texas CARES uses a convenience sample of Texans representing the four populations across the state.

For Phase I, on the day patients presented for their healthcare appointments, an FQHC healthcare team member offered adults 18-80 years of age literature on the Texas CARES Study and the Roche Elecsys® Anti-SARS-CoV-2 test, and invited them or their children (5-17 years of age) to enroll in the study. Participation was limited to two representatives from the same household between 5-80 years of age. Enrollment required contact information, demographic characteristics and informed consent for three blood draws over 6-12 months. Patients who consented to enroll in Texas CARES were provided a questionnaire collecting demographic information, employment, baseline medical conditions and comorbidities, prior COVID-19 tests and diagnoses, physician diagnosis of COVID-19 and other high-risk chronic illnesses such as type 2 diabetes, asthma and hypertension, COVID-19 symptoms and severity, and COVID-19 behavioral health^25^.

#### SARS Cov-2 Antibody Assay Roche Diagnostics

The primary outcome was a positive antibody assay qualitatively assessed using the Roche Elecsys® Anti-SARS-CoV-2 Immunoassay developed to detect antibodies to SARS-Cov-2. The Roche assay detects high-affinity antibodies to SARS-CoV-2 using a modified recombinant protein representing the nucleocapsid (N) antigen for the determination of SARS-CoV-2 antibodies. The assay relies on a double antigen sandwich (DAGS) format that enriches detection of higher affinity antibodies, which are more likely to be specific for SARS-CoV-2. The assay format is agnostic to the antibody isotype and can detect high affinity antibodies of all isotypes, it preferentially detects IgG antibodies since these are most likely to evolve to become high affinity. The nucleocapsid antigen is abundantly expressed and is a useful target for sensitive detection of virus-specific antibodies. These features provide an optimal combination of high specificity and sensitivity for the detection of immune exposure to SARS-CoV-2 in the general population, including in pregnant women and pediatric populations. The test has a published sensitivity of 99.82% sensitivity (95% CI: 99.69-99.91) and 99.91% specificity in diagnostic specimens (n=2861)^26^.

#### Questionnaire

A programmed questionnaire was designed to be completed in 10-15 minutes to capture demographic and clinical characteristics including comorbidities, prior COVID-19 virus testing, positivity, COVID-19 symptoms, previous antibody testing and mental health during the pandemic^27^. To help ensure validity, wherever possible, all questionnaire headers, questions, and response formats were harmonized to the PhenX Toolkit for COVID -19 and the BRFSS questionnaires. PhenX Toolkit items were reviewed for appropriateness, BRFSS and U.S. Census race/ethnicity questions were used. The self-administered questionnaire was designed to flow after completion of informed consent using a seamless webpage transition to ease respondent burden and maximize survey completion. All study materials, including the questionnaire, were available in both English and Spanish.

It was decided *a priori* that a survey weblink would be emailed and texted to those completing fewer than 50% of questions at their medical visit^29-30^. Those who did not respond by completing the survey received a phone call from a team member to collect the survey data. The survey completion percentage in our phase I study of 319 participants prior to the phone call was 96%, which is an indicator both of good validity and construction of our process for consent, survey completion and blood draw for antibody testing.

### Primary Outcomes and Statistical Analysis

The primary outcomes of the Phase I study included: 1) feasibility of implementation of the questionnaire and SARS-CoV-2 testing in a highly vulnerable population including children, and 2) estimation of Texas demographic and assay-adjusted cross-sectional seroprevalence based on antibody test results in these participants.

### Prevalence Estimation Methods

The SARS-CoV-2 cumulative prevalence was estimated from observed antibody reactivity using two sequential steps: (1) post-stratification weighting to standardize to the Texas population and (2) adjustment by antibody test sensitivity and specificity. First, crude observed seroprevalence was adjusted by age- and sex using weights derived from the U.S. census population projections for the state of Texas. Age in years was categorized into four categories: 18 to 34 years, 35 to 44 years, 45 to 54 years, and 55 years or greater [new reference]. Post-stratification weights were computed to standardize our sample to the greater Texas population according to the 2019 projected census; the weight was computed as a ratio of the proportion of a given level of a stratum in the census, divided by the equivalent proportion in the sample. An adjustment for the assay sensitivity (99.82%) and specificity (99.91%) was applied as per [Royal KD, 2019]. The full adjustment analysis was completed using SPSS version 27 and by hand. The weights are then applied to the individuals in our data set using standard survey weighting methods. Finally, to adjust for assay characteristics, the cumulative adjusted prevalence is computed as per Rosenberg et al.,^4^

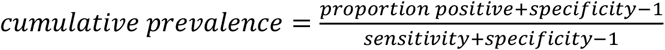

Estimates that are age and sex-standardized and adjusted for test characteristics are henceforth called “fully adjusted estimates.” “Crude estimates” refer to the observed seroprevalence estimates. Demographics, baseline variables, prior COVID-19 diagnoses and symptoms are also presented by crude antibody status.

## Results

### SARS-CoV-2 antibodies among total sample

Crude and adjusted SARS-CoV-2 antibody seropositivity are shown in Table 1. The crude antibody positivity prevalence in Phase I was 26.1% (80/307). The fully adjusted seroprevalence of the sample was 31.5%. Specifically, 41.1% of males and 21.9% of females were seropositive. For age categories, 33.5% of those 18-34; 24.4% of those 35-44; 33.2% of those 45-54; and 32.8% of those 55+ were seropositive.

**Table 1:** Seroprevalence rates by age and gender, standardized to Texas Census data and adjusted for Assay characteristics.

|  |  | Crude (observed) Antibody N and % |  |  |  | TX Census Weighted N and % |  |  |  |  | Adjusted for Assay (Spec=99.81%) |  |  |
| --- | --- | --- | --- | --- | --- | --- | --- | --- | --- | --- | --- | --- | --- |
|  |  | Antibody N |  | Antibody % |  |  | Weighted Antibody N |  | Weighted Antibody % |  | Adjusted Antibody Proportion |  |  |
|  | Phase I | Positive | Negative | Positive | Negative | Weighted | Positive | Negative | Positive | Negative | Positive | Estimated Infection experienced | Proportion of Estimated Infection experienced |
| Overall | 307 | 80 | 227 | 26.1% | 73.9% | 307 | 97 | 210 | 31.6% | 68.4% | 31.5% | 6795393 | 100% |
| Sex |  |  |  |  |  |  |  |  |  |  |  |  |  |
| Male | 63 | 26 | 37 | 41.3% | 58.7% | 155 | 62 | 89 | 41.1% | 58.9% | 41.0% | 4352534 | 64.1% |
| Female | 243 | 54 | 189 | 22.2% | 77.8% | 152 | 35 | 121 | 22.4% | 77.6% | 22.3% | 2444240 | 36.0% |
| Age group |  |  |  |  |  |  |  |  |  |  |  |  |  |
| 18-34 | 90 | 24 | 66 | 26.7% | 73.3% | 98 | 33 | 65 | 33.7% | 66.3% | 33.5% | 2378518 | 35.0% |
| 35-44 | 69 | 14 | 55 | 20.3% | 79.7% | 58 | 14 | 43 | 24.6% | 75.4% | 24.4% | 962657 | 14.2% |
| 45-54 | 74 | 23 | 51 | 31.1% | 68.9% | 76 | 25 | 50 | 33.3% | 66.7% | 33.2% | 1180189 | 17.4% |
| 55+ | 74 | 19 | 55 | 25.7% | 74.3% | 76 | 24 | 51 | 32.0% | 68.0% | 31.9% | 2233957 | 32.9% |

### Demographic and clinical correlates of seropositivity

Demographic and clinical characteristics, by SARS-CoV-2 antibody seropositivity are presented for the total Phase I sample, FQHC clinical staff, and FQHC patient population in Tables 2a-c. As shown in Table 2a, 17.7% (14/79) FQHC employees tested were positive and 27.9% (57/204) of FQHC patients were positive. The mean age of the entire sample (N=319) was 43.7 (SD=13.5). The group was primarily female (79%, n=252), white (95.3%, n=286), and of Hispanic ethnicity (81.7%, n=255), with 8.7% (n=25) having some high school or less and 19.8% (n=57) having an advanced professional or academic degree. A total of 78% (n=221)] was employed full-time and 79% (n=228) reported having some type of health insurance. The clinical characteristics of the sample indicate that 27.7% (n=78) were overweight and 59.9% (n=169) were obese with the mean BMI = 32.6 (SD=7.6). The majority of participants reported not using tobacco products in the past two weeks (88.7%, n=260) and did not report use of vaping products in the past two weeks (96.8%, n=272).

**Table 2a:** Demographics and clinical characteristics, TX CARES, all phase 1 participants, 2020.

| Demographics | Overall<br>(n=319)* | SARS-CoV2 Antibody Status |  |
| --- | --- | --- | --- |
|  |  | Positive (n=80) | Negative (n=227) |
|  | N (%) | N (%) | N (%) |
| Cohort (n=291) |  |  |  |
| FQHC Employee | 82 (25.7) | 14 (17.5) | 65 (28.6) |
| FQHC Patient | 209 (65.5) | 57 (71.3) | 147 (64.8) |
| Missing | 28 (8.8) | 9 (11.3) | 15 (6.6) |
| Gender (n=319) |  |  |  |
| Male | 67 (21.0) | 26 (32.5) | 38 (16.7) |
| Female | 252 (79.0) | 54 (67.5) | 189 (83.3) |
| Missing | 0 (0.0) | 0 (0.0) | 0 (0.0) |
| Race (n=300) |  |  |  |
| White | 286 (89.7) | 73 (91.2) | 202 (89.0) |
| Black/African American | 5 (1.6) | 0 (0.0) | 5 (2.2) |
| Asian | 3 (0.9) | 1 (1.2) | 2 (0.9) |
| Hawaiian or Other<br>Pacific Islander | 1 (0.3) | 0 (0.0) | 1 (0.4) |
| American Indian or<br>Alaska Native | 2 (0.6) | 0 (0.0) | 1 (0.4) |
| Multi-racial | 3 (0.9) | 1 (1.2) | 2 (0.9) |
| Missing | 19 (6.0) | 5 (6.2) | 14 (6.2) |
| Hispanic Ethnicity (n= 312) |  |  |  |
| Yes | 255 (79.9) | 75 (93.8) | 171 (75.3) |
| No | 57 (17.9) | 3 (3.8) | 51 (22.5) |
| Missing | 7 (2.2) | 2 (2.5) | 5 (2.2) |
| Education (n= 288) |  |  |  |
| Less than high school | 12 (3.8) | 6 (7.5) | 6 (2.6) |
| Some high school | 13 (4.1) | 4 (5.0) | 9 (4.0) |
| High school graduate/GED | 78 (24.5) | 27 (33.8) | 48 (21.1) |
| Some college, no degree | 68 (21.3) | 20 (25.0) | 48 (21.1) |
| Two- or four-year college degree | 60 (18.8) | 12 (15.0) | 46 (20.3) |
| Advanced professional or academic degree | 57 (17.9) | 4 (5.0) | 50 (22.0) |
| Missing | 31 (9.7) | 7 (8.8) | 20 (8.8) |
| Employment (n= 283) |  |  |  |
| Full-time | 221 (69.3) | 39 (48.8) | 175 (77.1) |
| Part-time | 12 (3.8) | 6 (7.5) | 6 (2.6) |
| Unemployed | 38 (11.9) | 19 (23.8) | 18 (7.9) |
| Other | 12 (3.8) | 5 (6.2) | 7 (3.1) |
| Missing | 36 (11.3) | 11 (13.8) | 21 (9.3) |
| Has Insurance (n= 289) |  |  |  |
| Yes | 228 (71.5) | 40 (50.0) | 181 (79.7) |
| No | 61 (19.1) | 30 (47.5) | 30 (13.2) |
| Missing | 30 (9.4) | 10 (12.5) | 16 (7.0) |
| BMI, categorical (n= 282) |  |  |  |
| Underweight | 3 (0.9) | 2 (2.5) | 1 (0.4) |
| Normal | 32 (10.0) | 6 (7.5) | 23 (10.1) |
| Overweight | 78 (24.5) | 21 (26.2) | 56 (24.7) |
| Obese | 169 (53.0) | 41 (51.2) | 124 (54.6) |
| Missing | 37 (11.6) | 10 (12.5) | 23 (10.1) |
| Frequency of Smoking or Use of<br>Other Tobacco Products in Past<br>Two Weeks (n= 293) |  |  |  |
| Not at all | 260 (81.5) | 67 (83.8) | 186 (81.9) |
| Rarely | 8 (2.5) | 2 (2.5) | 6 (2.6) |
| Once a day | 9 (2.8) | 3 (3.8) | 5 (2.2) |
| More than once a day | 16 (5.0) | 1 (1.2) | 15 (6.6) |
| Missing | 26 (8.2) | 7 (8.8) | 15 (6.6) |

| Frequency of Using Vaping<br>Products in Past Two Weeks (n= 281) |  |  |  |
| --- | --- | --- | --- |
| Not at all | 272 (85.3) | 69 (86.2) | 195 (85.9) |
| Rarely | 6 (1.9) | 0 (0.0) | 6 (2.6) |
| Once a day | 0 (0.0) | 0 (0.0) | 0 (0.0) |
| More than once a day | 3 (0.9) | 1 (1.2) | 2 (0.9) |
| Missing | 38 (11.9) | 10 (12.5) | 24 (10.6) |
|  | **Mean (±SD)** | **Mean (±SD)** | **Mean (±SD)** |
| Age, years (n= 317;<br>missing = 2) | 43.7 (13.5) | 43.3 (13.8) | 43.6 (13.4) |
| Height, inches (n= 287;<br>missing = 32) | 64.2 (3.8) | 64.6 (4.5) | 64.0 (3.6) |
| Weight, pounds (n= 288;<br>missing = 31) | 191.4 (46.9) | 188.1 (43.5) | 192.0 (47.3) |
| BMI, continuous (n= 281;<br>missing = 38) | 32.6 (7.6) | 31.6 (6.8) | 33.0 (7.8) |

\*12 individuals completed a survey, but did not complete an antibody test

**Table 2b:** Demographics and clinical characteristics, TX CARES, phase 1 clinic employees, 2020.

| Demographics | Overall<br>(n=82)* | SARS-CoV2 Antibody Status |  |
| --- | --- | --- | --- |
|  |  | Positive (n=14) | Negative (n=65) |
|  | N (%) | N (%) | N (%) |
| Gender (n=82) |  |  |  |
| Male | 10 (12.2) | 2 (14.3) | 7 (10.8) |
| Female | 72 (87.8) | 12 (85.7) | 58 (89.2) |
| Missing | 0 (0.0) | 0 (0.0) | 0 (0.0) |
| Race (n=78) |  |  |  |
| White | 73 (89.0) | 12 (85.7) | 59 (90.8) |
| Black/African American | 2 (2.4) | 0 (0.0) | 2 (3.1) |
| Asian | 2 (2.4) | 0 (0.0) | 2 (3.1) |
| Hawaiian or Other<br>Pacific Islander | 0 (0.0) | 0 (0.0) | 0 (0.0) |
| American Indian or<br>Alaska Native | 1 (1.2) | 0 (0.0) | 0 (0.0) |
| Missing | 4 (4.9) | 2 (14.3) | 2 (3.1) |
| Hispanic Ethnicity (n=81) |  |  |  |
| Yes | 61 (74.4) | 14 (100.0) | 45 (69.2) |
| No | 20 (24.4) | 0 (0.0) | 19 (29.2) |
| Missing | 1 (1.2) | 0 (0.0) | 1 (1.5) |
| Education (n=80) |  |  |  |
| Less than high school | 0 (0.0) | 0 (0.0) | 0 (0.0) |
| Some high school | 1 (1.2) | 1 (7.1) | 0 (0.0) |
| High school graduate/GED | 20 (24.4) | 4 (28.6) | 15 (23.1) |
| Some college, no degree | 27 (32.9) | 4 (28.6) | 23 (35.4) |
| Two- or four-year college degree | 14 (17.1) | 4 (28.6) | 9 (13.8) |
| Advanced professional or academic degree | 18 (22.0) | 1 (7.1) | 16 (24.6) |
| Missing | 2 (2.4) | 0 (0.0) | 2 (3.1) |
| Employment (n=80) |  |  |  |
| Full-time | 79 (96.3) | 13 (92.9) | 63 (96.9) |
| Part-time | 0 (0.0) | 0 (0.0) | 0 (0.0) |
| Unemployed | 0 (0.0) | 0 (0.0) | 0 (0.0) |
| Other | 1 (1.2) | 1 (7.1) | 0 (0.0) |
| Missing | 2 (2.4) | 0 (0.0) | 2 (3.1) |
| Has Insurance (n=82) |  |  |  |
| Yes | 73 (89.0) | 11 (78.6) | 60 (92.3) |
| No | 9 (11.0) | 3 (21.4) | 5 (7.7) |
| Missing | 0 (0.0) | 0 (0.0) | 0 (0.0) |
| BMI, categorical (n=79) |  |  |  |
| Underweight | 1 (1.2) | 1 (7.1) | 0 (0.0) |
| Normal | 10 (12.2) | 2 (14.3) | 7 (10.8) |
| Overweight | 12 (14.6) | 2 (14.3) | 9 (13.8) |
| Obese | 56 (68.3) | 8 (57.1) | 47 (72.3) |
| Missing | 3 (3.7) | 1 (7.1) | 2 (3.1) |
| Frequency of Smoking or Use of<br>Other Tobacco Products in Past<br>Two Weeks (n=82) |  |  |  |
| Not at all | 71 (86.6) | 13 (92.9) | 56 (86.2) |
| Rarely | 1 (1.2) | 0 (0.0) | 1 (1.5) |
| Once a day | 4 (4.9) | 1 (7.1) | 2 (3.1) |
| More than once a day | 6 (7.3) | 0 (0.0) | 6 (9.2) |
| Missing | 0 (0.0) | 0 (0.0) | 0 (0.0) |
| Frequency of Using Vaping<br>Products in Past Two Weeks (n=77) |  |  |  |
| Not at all | 74 (90.2) | 12 (85.7) | 59 (90.8) |
| Rarely | 1 (1.2) | 0 (0.0) | 1 (1.5) |
| Once a day | 0 (0.0) | 0 (0.0) | 0 (0.0) |
| More than once a day | 2 (2.4) | 0 (0.0) | 2 (3.1) |
| Missing | 5 (6.1) | 2 (14.3) | 3 (4.6) |
|  | **Mean (±SD)** | **Mean (±SD)** | **Mean (±SD)** |
| Age, years (n=82;<br>missing = 0) | 40.1 (11.8) | 49.29 (9.2) | 37.7 (11.2) |
| Height, inches (n=81;<br>missing = 1) | 64.0 (3.6) | 63.50 (3.4) | 64.1 (3.6) |
| Weight, pounds (n=80;<br>missing = 2) | 198.6 (46.5) | 174.23 (43.6) | 204.0 (46.2) |
| BMI, continuous (n=79;<br>missing = 3) | 34.1 (7.7) | 30.25 (6.6) | 35.0 (7.8) |

**\*3 individuals completed a survey, but did not complete an antibody test**

**Table 2c:** Demographics and clinical characteristics, TX CARES, phase 1 clinic patients, 2020.

| Demographics | Overall<br>(n=209)* | SARS-CoV2 Antibody Status |  |
| --- | --- | --- | --- |
|  |  | Positive (n=57) | Negative (n=147) |
|  | N (%) | N (%) | N (%) |
| Gender (n=209) |  |  |  |
| Male | 53 (25.4) | 23 (40.4) | 29 (19.7) |
| Female | 156 (74.6) | 34 (59.6) | 118 (80.3) |
| Missing | 0 (0.0) | 0 (0.0) | 0 (0.0) |
| Race (n=194) |  |  |  |
| White | 185 (88.5) | 52 (91.2) | 128 (87.1) |
| Black/African American | 3 (1.4) | 0 (0.0) | 3 (2.0) |
| Asian | 1 (0.5) | 1 (1.8) | 0 (0.0) |
| Hawaiian or Other<br>Pacific Islander | 1 (0.5) | 0 (0.0) | 1 (0.7) |
| American Indian or<br>Alaska Native | 1 (0.5) | 0 (0.0) | 1 (0.7) |
| Multi-racial | 3 (1.4) | 1 (1.8) | 2 (1.4) |
| Missing | 15 (7.2) | 3 (5.3) | 12 (8.2) |
| Hispanic Ethnicity (n=204) |  |  |  |
| Yes | 170 (81.3) | 52 (91.2) | 114 (77.6) |
| No | 34 (16.3) | 3 (5.3) | 30 (20.4) |
| Missing | 5 (2.4) | 2 (3.5) | 3 (2.0) |
| Education (n=206) |  |  |  |
| Less than high school | 12 (5.7) | 6 (10.5) | 6 (4.1) |
| Some high school | 11 (5.3) | 2 (3.5) | 9 (6.1) |
| High school graduate/GED | 57 (27.3) | 22 (38.6) | 33 (22.4) |
| Some college, no degree | 41 (19.6) | 16 (28.1) | 25 (17.0) |
| Two- or four-year college degree | 46 (22.0) | 8 (14.0) | 37 (25.2) |
| Advanced professional or academic degree | 39 (18.7) | 3 (5.3) | 34 (23.1) |
| Missing | 3 (1.4) | 0 (0.0) | 3 (2.0) |
| Employment (n=202) |  |  |  |
| Full-time | 141 (67.5) | 25 (43.9) | 112 (76.2) |
| Part-time | 12 (5.7) | 6 (10.5) | 6 (4.1) |
| Unemployed | 38 (18.2) | 19 (33.3) | 18 (12.2) |
| Other | 11 (5.3) | 4 (7.0) | 7 (4.8) |
| Missing | 7 (3.3) | 3 (5.3) | 4(2.7) |
| Has Insurance (n=206) |  |  |  |
| Yes | 154 (73.7) | 28 (49.1) | 121 (82.3) |
| No | 52 (24.9) | 27 (47.4) | 25 (17.0) |
| Missing | 3 (1.4) | 2 (3.5) | 1 (0.7) |
| BMI, categorical (n=199) |  |  |  |
| Underweight | 2 (1.0) | 1 (1.8) | 1 (0.7) |
| Normal | 22 (10.5) | 4 (7.0) | 16 (10.9) |
| Overweight | 66 (31.6) | 19 (33.3) | 47 (32.0) |
| Obese | 111 (53.1) | 31 (54.4) | 77 (52.4) |
| Missing | 8 (3.8) | 2 (3.5) | 6 (4.1) |
| Frequency of Smoking or Use of<br>Other Tobacco Products in Past<br>Two Weeks (n=209) |  |  |  |
| Not at all | 187 (89.5) | 52 (91.2) | 130 (88.4) |
| Rarely | 7 (3.3) | 2 (3.5) | 5 (3.4) |
| Once a day | 5 (2.4) | 2 (3.5) | 3 (2.0) |
| More than once a day | 10 (4.8) | 1 (1.8) | 9 (6.1) |
| Missing | 0 (0.0) | 0 (0.0) | 0 (0.0) |
| Frequency of Using Vaping<br>Products in Past Two Weeks (n=202) |  |  |  |
| Not at all | 196 (93.8) | 55 (96.5) | 136 (92.5) |
| Rarely | 5 (2.4) | 0 (0.0) | 5 (3.4) |
| Once a day | 0 (0.0) | 0 (0.0) | 0 (0.0) |
| More than once a day | 1 (0.5) | 1 (1.8) | 0 (0.0) |
| Missing | 7 (3.3) | 1 (1.8) | 6 (4.1) |
|  | **Mean ±SD)** | **Mean (±SD)** | **Mean (±SD)** |
| Age, years (n=209;<br>missing = 0) | 45.1(13.8) | 41.8 (14.15) | 46.0 (13.6) |
| Height, inches (n=203;<br>missing = 6) | 64.3 (4.0) | 65.0 (4.73) | 63.9 (3.7) |
| Weight, pounds (n=205;<br>missing = 4) | 188.5 (47.2) | 190.9 (43.75) | 186.6 (47.0) |
| BMI, continuous (n=201;<br>missing = 8) | 32.4 (7.8) | 31.7 (6.89) | 32.1 (7.6) |

**\*5 individuals completed a survey, but did not complete an antibody test**

### SARS-CoV-2 symptoms and previous diagnoses

In Table 3a, of those 80 people with a positive SARS-CoV-2 antibody test 78.9% (56/71) reported having had at least one symptom of COVID-19. Of those 227 who were negative, 38% (71/186) reported presence of COVID-19 symptoms. More than half (53.1%, 154/290) of the participants reported having had a previous COVID-19 test. Of the154, 152 responded whether that test was positive or negative: 61/152 indicated it was positive (40.1%). In this sample,42.2% (89/211) of those negative for the antibody test reported having had a COVID-19 test.

**Table 3a:** SARS-CoV2 Symptoms and Previous Diagnosis, TX CARES, all phase 1 participants, 2020.

| Previous COVID-19<br>Diagnosis/Symptoms | Overall<br>(n=319)* | SARS-CoV2 Antibody Status |  |
| --- | --- | --- | --- |
|  |  | Positive<br>(n=80) | Negative<br>(n=227) |
|  | N (%) | N (%) | N (%) |
| Any COVID-19 Symptoms (n= 265) |  |  |  |
| Yes | 130 (40.8) | 56 (70.0) | 71 (31.3) |
| No | 135 (42.3) | 15 (18.8) | 115 (50.7) |
| Missing | 54 (16.9) | 9 (11.3) | 41 (18.1) |
| Previous COVID-19 Test (n= 290) |  |  |  |
| Yes | 154 (48.3) | 62 (77.5) | 89 (39.2) |
| No | 136 (42.6) | 10 (12.5) | 122 (53.7) |
| Missing | 29 (9.1) | 8 (10.0) | 16 (7.0) |
| Previous Positive COVID-19<br>Test Result (n= 152) |  |  |  |
| Yes | 61 (19.1) | 55 (68.8) | 6 (2.6) |
| No | 91 (28.5) | 6 (7.5) | 82 (36.1) |
| Missing | 167 (52.4) | 19 (23.8) | 139 (61.2) |
| Diagnosed with COVID-19 by<br>Health Professional Without Test (n= 290) |  |  |  |
| Yes | 11 (3.4) | 7 (8.8) | 3 (1.3) |
| No | 279 (87.5) | 65 (81.2) | 207 (91.2) |
| Missing | 29 (9.1) | 4 (33.3) | 17 (7.5) |
|  | **Median (IQR)** | **Median (IQR)** | **Median (IQR)** |
| **Symptoms\*\*** |  |  |  |
| Fever or Chills (n= 68) | 3.00 (2.00 - 4.00) | 3.00 (2.00 - 4.00) | 3.00 (2.50 - 4.00) |
| Missing n(%) | 253 (79.3) | 51 (63.8) | 192 (84.6) |
| Cough (n= 71) | 3.00 (2.00 - 4.00) | 3.00 (2.00 - 4.00) | 3.00 (2.00 - 4.00) |
| Missing n(%) | 249 (78.1) | 48 (60.0) | 190 (83.7) |
| Shortness of<br>Breath/Difficulty<br>Breathing (n= 53) | 3.00 (2.00 - 4.00) | 3.50 (3.00 - 4.75) | 3.00 (2.00 - 4.00) |
| Missing n(%) | 266 (83.4) | 50 (62.5) | 206 (90.7) |
| Fatigue(n= 79) | 4.00 (3.00 - 4.00) | 4.00 (3.00 - 5.00) | 3.00 (3.00 - 4.00) |
| Missing n(%) | 242 (75.9) | 47 (58.8) | 185 (81.5) |
| Muscle or Body Aches<br>(n=72) | 3.00 (3.00 - 4.00) | 4.00 (3.00 - 4.75) | 3.00 (3.00 - 4.00) |
| Missing n(%) | 248 (77.7) | 46 (57.5) | 192 (84.6) |
| Headaches (n=83) | 4.00 (3.00 - 5.00) | 4.00 (3.00 - 5.00) | 3.00 (3.00 - 4.00) |
| Missing n(%) | 237 (74.3) | 46 (57.5) | 181 (79.7) |
| Congestion or Runny Nose (n= 63) | 3.00 (2.00 - 4.00) | 3.00 (2.00 - 4.00) | 3.00 (2.00 - 4.00) |
| Missing n(%) | 257 (80.6) | 55 (68.8) | 190 (83.7) |
| Diarrhea (n=44) | 3.00 (2.00 - 3.00) | 3.00 (2.00 - 4.00) | 3.00 (2.00 - 3.00) |
| Missing n(%) | 276 (86.5) | 60 (75.0) | 204 (89.9) |
| Nausea or Vomiting (n=37) | 3.00 (2.00 - 4.00) | 3.00 (2.00 - 5.00) | 3.00 (2.00 - 4.00) |
| Missing n(%) | 282 (88.4) | 63 (78.8) | 207 (91.2) |
| New Loss of Taste or Smell (n=44) | 5.00 (4.00 - 5.00) | 5.00 (4.00 - 5.00) | 4.00 (3.50 - 5.00) |
| Missing n(%) | 276 (86.5) | 45 (56.3) | 220 (96.9) |
| Sore Throat (n=68) | 3.00 (2.00 - 4.00) | 3.00 (2.00 - 4.00) | 3.00 (2.00 - 3.25) |
| Missing n(%) | 253 (79.3) | 55 (68.8) | 187 (82.4) |

**\* 12 individuals completed a survey, but did not complete an antibody test**
**\*\*Symptom severity based on a scale from 1 to 5 with 1 being minimal and 5 being severe**

Of the 61 respondents with a prior positive COVID-19 test, 55 (90.2%) had antibodies and 6 (9.8%) did not have antibodies. Of those diagnosed with COVID-19 by a health professional without a test, 7 (70.0%) had a positive antibody test and 3 (30%) had a negative antibody test result. The most commonly reported symptoms in the sample positive for SARS-CoV-2 antibodies were new loss of taste or smell, fatigue, muscle or body aches, and headaches. SARS-CoV-2 symptoms and previous diagnoses by clinic employee and clinic patient are reported in Tables 3b and 3c.

**Table 3b:** SARS-CoV2 Symptoms and Previous Diagnosis, TX CARES, phase 1 clinic employees, 2020.

| Previous COVID-19<br>Diagnosis/Symptoms | Overall<br>(n=82)* | SARS-CoV2 Antibody Status |  |
| --- | --- | --- | --- |
|  |  | Positive<br>(n=14) | Negative<br>(n=65) |
|  | N (%) | N (%) | N (%) |
| Any COVID-19 Symptoms (n=76) |  |  |  |
| Yes | 39 (47.6) | 12 (85.7) | 26 (40.0) |
| No | 37 (45.1) | 1 (7.1) | 33 (50.8) |
| Missing | 6 (7.3) | 1 (7.1) | 6 (9.2) |
| Previous COVID-19 Test (n=81) |  |  |  |
| Yes | 48 (58.5) | 13 (92.9) | 33 (50.8) |
| No | 33 (40.2) | 1 (7.1) | 31 (47.7) |
| Missing | 1 (1.2) | 0 (0.0) | 1 (1.5) |
| Previous Positive COVID-19<br>Test Result (n=48) |  |  |  |
| Yes | 14 (17.1) | 13 (92.9) | 1 (1.5) |
| No | 34 (41.5) | 0 (0.0) | 32 (49.2) |
| Missing | 34 (41.5) | 1 (7.1) | 32 (49.2) |
| Diagnosed with COVID-19 by<br>Health Professional Without Test (n=81) |  |  |  |
| Yes | 4 (4.9) | 3 (21.4) | 1 (1.5) |
| No | 77 (93.9) | 11 (78.6) | 63 (96.9) |
| Missing | 1 (1.2) | 0 (0.0) | 1 (1.5) |
|  | **Median (IQR)** | **Median (IQR)** | **Median (IQR)** |
| **Symptoms\*\*** |  |  |  |
| Fever or Chills (n=28) | 3.00 (2.25 - 4.00) | 4.00 (3.50 - 4.00) | 3.00 (2.00 - 3.00) |
| Missing n(%) | 64 (78.0) | 7 (50.0) | 54 (83.1) |
| Cough (n=27) | 3.00 (2.00 - 4.00) | 4.00 (3.50 - 5.00) | 3.00 (2.00 - 3.00) |
| Missing n(%) | 65 (79.3) | 7 (50.0) | 55 (84.6) |
| Shortness of<br>Breath/Difficulty<br>Breathing (n=22) | 3.00 (2.25 - 4.00) | 4.00 (3.00 - 4.00) | 2.00 (1.00 - 2.00) |
| Missing n(%) | 68 (82.9) | 5 (35.7) | 60 (70.6) |
| Fatigue(n=34) | 4.00 (3.00 - 4.00) | 4.50 (3.75 - 5.00) | 3.00 (3.00 - 4.00) |
| Missing n(%) | 57 (69.5) | 6 (42.9) | 48 (73.8) |
| Muscle or Body Aches<br>(n=29) | 3.00 (3.00 - 4.00) | 4.00 (3.00 - 5.00) | 3.00 (2.75 - 4.00) |
| Missing n(%) | 61 (74.4) | 5 (35.7) | 53 (81.5) |
| Headaches (n=32) | 4.00 (3.00 - 5.00) | 5.00 (4.00 - 5.00) | 3.00 (3.00 - 4.00) |
| Missing n(%) | 59 (72.0) | 6 (42.9) | 50 (76.9) |
| Congestion or Runny Nose (n=29) | 3.00 (3.00 - 4.50) | 4.00 (2.50 - 5.00) | 3.00 (3.00 - 4.00) |
| Missing n(%) | 63 (76.8) | 7 (50.0) | 53 (81.5) |
| Diarrhea (n=25) | 2.50 (2.00 - 3.00) | 2.50 (2.00 - 3.50) | 2.50 (2.00 - 3.00) |
| Missing n(%) | 70 (85.4) | 10 (71.4) | 57 (87.7) |
| Nausea or Vomiting (n=23) | 3.00 (2.00 - 4.00) | 3.50 (2.75 - 4.25) | 2.50 (1.25 - 3.75) |
| Missing n(%) | 72 (87.8) | 10 (71.4) | 59 (90.8) |
| New Loss of Taste or Smell (n=19) | 5.00 (4.25 - 5.00) | 5.00 (4.50 - 5.00) | 4.50 (4.25 - 4.75) |
| Missing n(%) | 72 (87.8) | 6 (42.9) | 63 (96.9) |
| Sore Throat (n=33) | 3.00 (2.00 - 3.50) | 4.00 (3.00 - 4.00) | 3.00 (2.00 - 3.00) |
| Missing n(%) | 59 (72.0) | 7 (50.0) | 49 (75.4) |

**\* 3 individuals completed this part of the survey, but did not complete an antibody test**
**\*\*Symptom severity based on a scale from 1 to 5 with 1 being minimal and 5 being severe**

**Table 3c:**
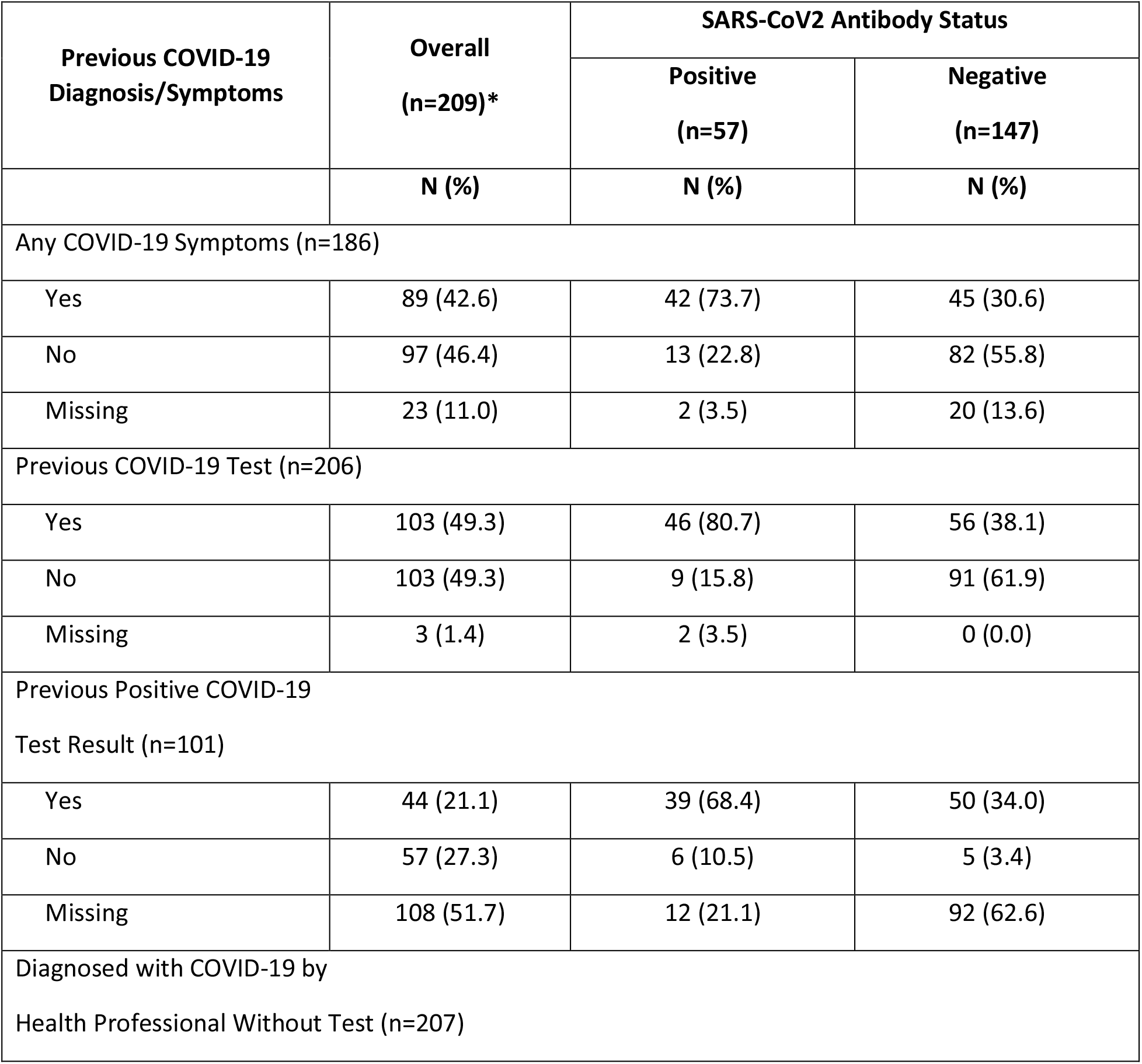
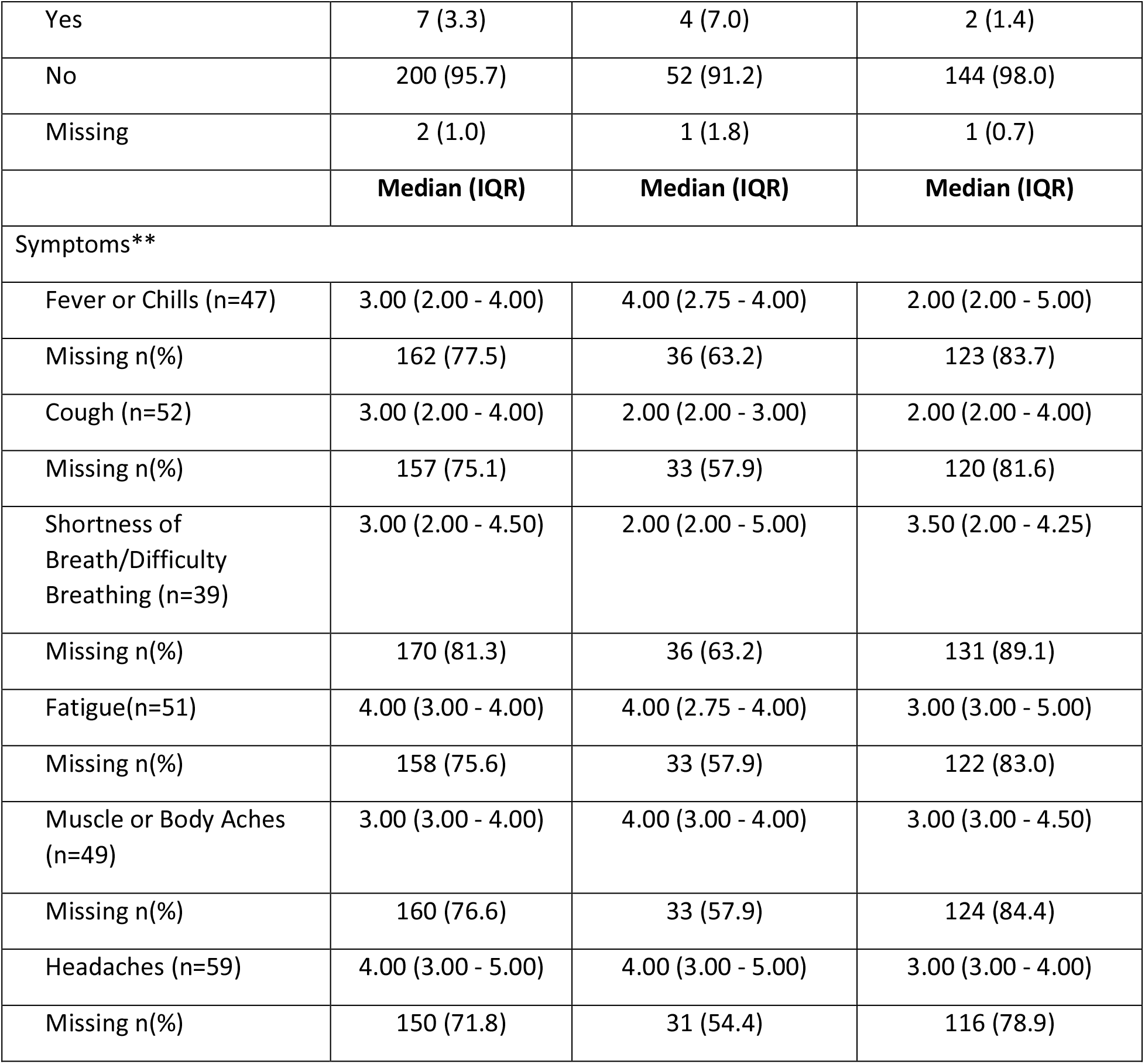
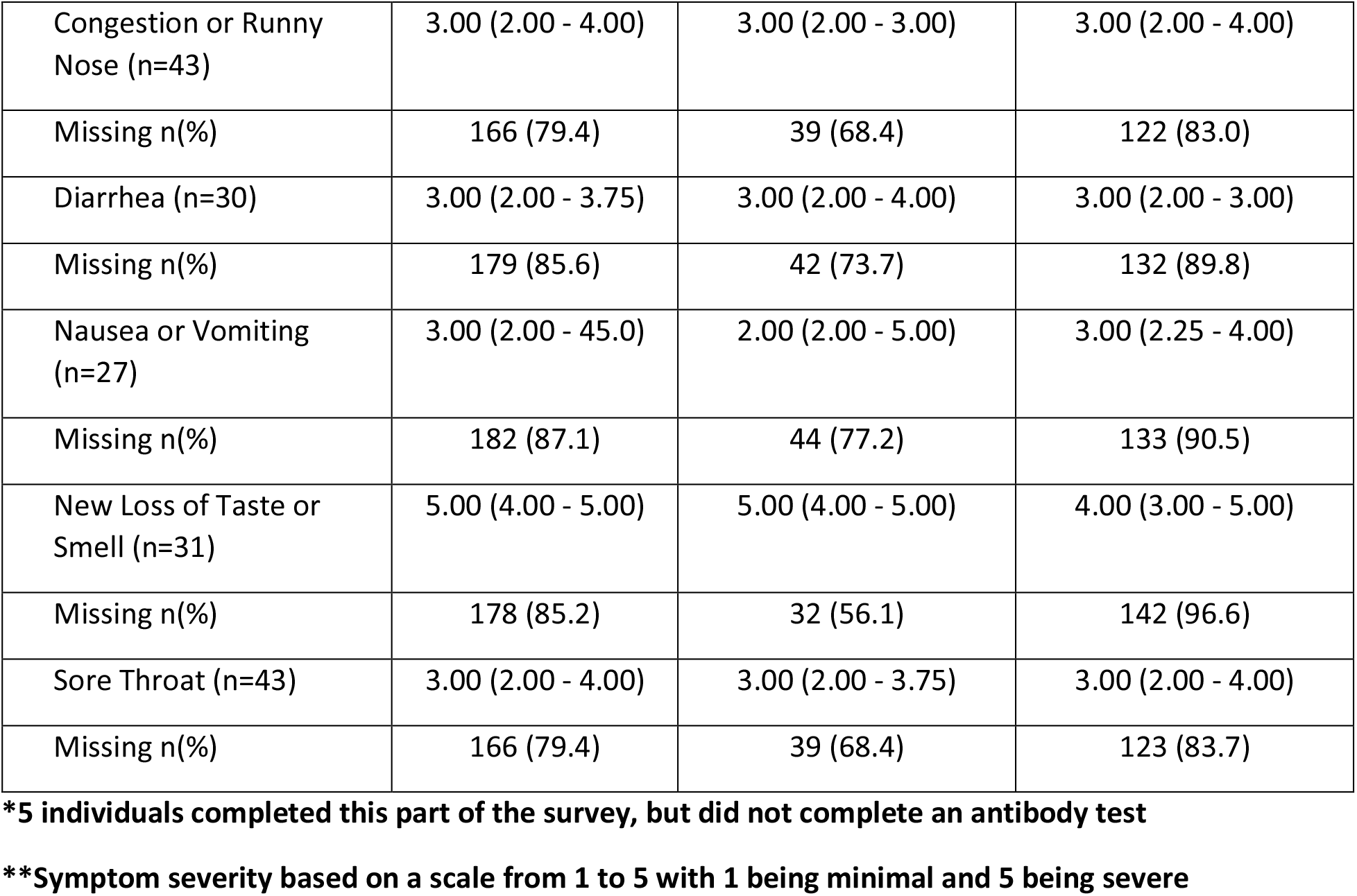
SARS-CoV2 Symptoms and Previous Diagnosis, TX CARES, phase 1 clinic patients, 2020.

### Presence of chronic diseases in the Texas CARES Phase I sample

From Table 4c, those with a positive SARS-CoV-2 antibody test were most likely to report having the following chronic diseases: hypertension (21/68, 30.9%), diabetes (18/68, 26.5%), asthma (18/68, 26.5%), and obesity (14/68, 20.6%). Chronic disease reports by clinic employee and clinic patients are presented in Tables 4b and 4c.

**Table 4a:**
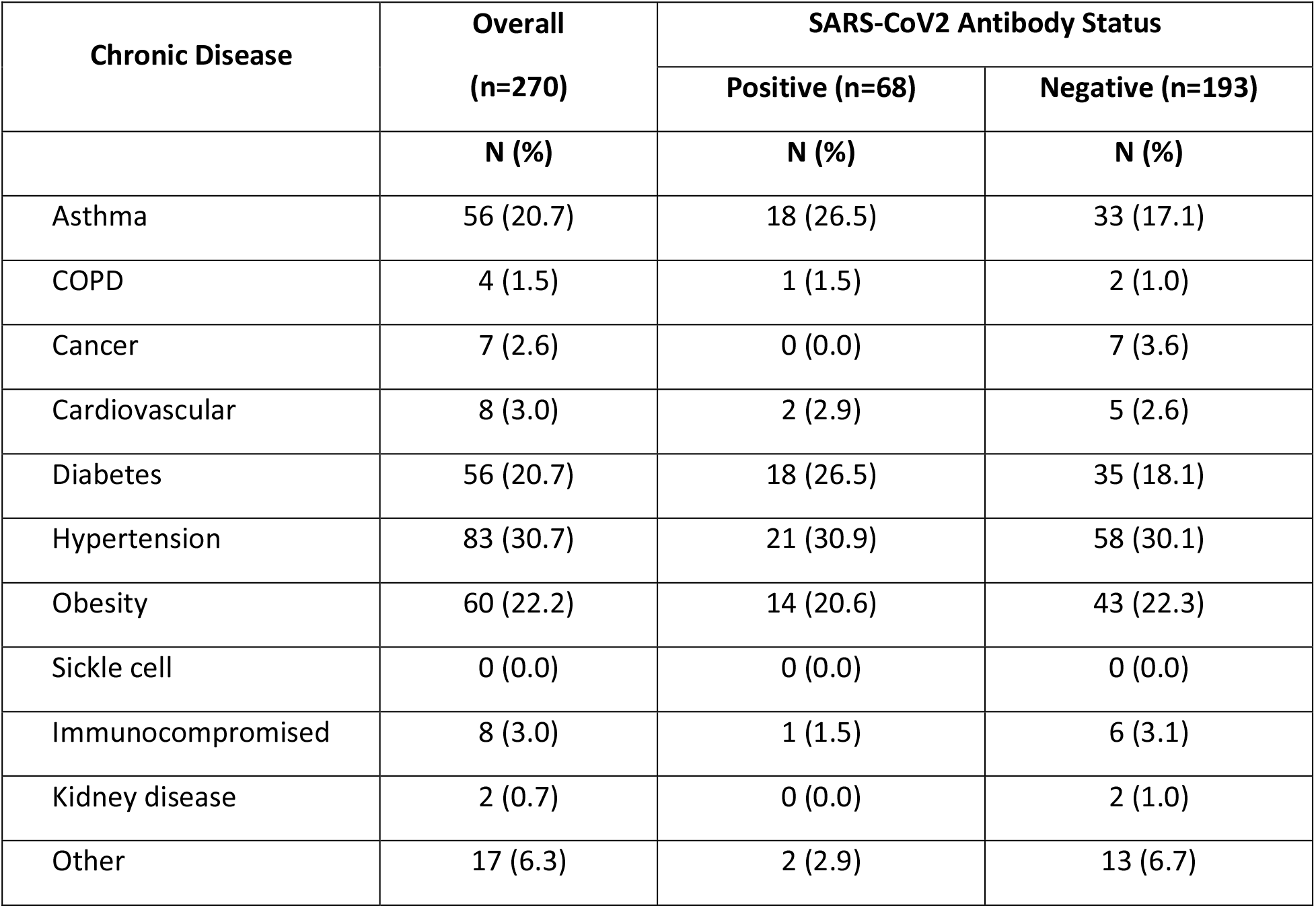
Chronic Diseases, TX CARES, all phase 1 participants, 2020.

| Chronic Disease | Overall<br>(n=270) | SARS-CoV2 Antibody Status |  |
| --- | --- | --- | --- |
|  |  | Positive (n=68) | Negative (n=193) |
|  | N (%) | N (%) | N (%) |
| Asthma | 56 (20.7) | 18 (26.5) | 33 (17.1) |
| COPD | 4 (1.5) | 1 (1.5) | 2 (1.0) |
| Cancer | 7 (2.6) | 0 (0.0) | 7 (3.6) |
| Cardiovascular | 8 (3.0) | 2 (2.9) | 5 (2.6) |
| Diabetes | 56 (20.7) | 18 (26.5) | 35 (18.1) |
| Hypertension | 83 (30.7) | 21 (30.9) | 58 (30.1) |
| Obesity | 60 (22.2) | 14 (20.6) | 43 (22.3) |
| Sickle cell | 0 (0.0) | 0 (0.0) | 0 (0.0) |
| Immunocompromised | 8 (3.0) | 1 (1.5) | 6 (3.1) |
| Kidney disease | 2 (0.7) | 0 (0.0) | 2 (1.0) |
| Other | 17 (6.3) | 2 (2.9) | 13 (6.7) |

**Table 4b:** Chronic Diseases, TX CARES, phase 1 clinic employees, 2020.

| Chronic Disease | Overall<br>(n= 69) | SARS-CoV2 Antibody Status |  |
| --- | --- | --- | --- |
|  |  | Positive (n= 12) | Negative (n=53) |
|  | N (%) | N (%) | N (%) |
| Asthma | 13 (18.8) | 3 (25.0) | 7 (13.2) |
| COPD | 1 (1.4) | 0 (0.0) | 1 (1.9) |
| Cancer | 0 (0.0) | 0 (0.0) | 0 (0.0) |
| Cardiovascular | 2 (2.9) | 1 (8.3) | 1 (1.9) |
| Diabetes | 8 (11.6) | 2 (16.7) | 5 (9.4) |
| Hypertension | 15 (21.7) | 3 (25.0) | 11 (20.8) |
| Obesity | 25 (36.2) | 4 (33.3) | 21 (39.6) |
| Sickle cell | 0 (0.0) | 0 (0.0) | (0.0) |
| Immunocompromised | 2 (2.9) | 0 (0.0) | 2 (3.8) |
| Kidney disease | 1 (1.4) | 0 (0.0) | 1 (1.9) |
| Other | 2 (2.9) | 0 (0.0) | 2 (3.8) |

**Table 4c:** Chronic Diseases, TX CARES, phase 1 clinic patients, 2020.

| Chronic Disease | Overall<br>(n= 199) | SARS-CoV2 Antibody Status |  |
| --- | --- | --- | --- |
|  |  | Positive (n=54) | Negative (n=140) |
|  | N (%) | N (%) | N (%) |
| Asthma | 43 (21.6) | 15 (27.8) | 26 (18.6) |
| COPD | 3 (1.5) | 1 (1.9) | 1 (0.7) |
| Cancer | 7 (3.5) | 0 (0.0) | 7 (5.0) |
| Cardiovascular | 6 (3.0) | 1 (1.9) | 4 (2.9) |
| Diabetes | 48 (24.1) | 16 (29.6) | 30 (21.4) |
| Hypertension | 68 (34.2) | 18 (33.3) | 47 (33.6) |
| Obesity | 35 (17.6) | 10 (18.5) | 22 (15.7) |
| Sickle cell | 0 (0.0) | 0 (0.0) | 0 (0.0) |
| Immunocompromised | 6 (3.0) | 1 (1.9) | 4 (2.9) |
| Kidney disease | 1 (0.5) | 0 (0.0) | 1 (0.7) |
| Other | 15 (7.5) | 2 (3.7) | 11 (7.9) |

## Discussion

In this study we enrolled and analyzed 319 participants, demonstrating a high survey and antibody test completion rate, and ability to implement a questionnaire and SARS-CoV-2 antibody testing within FQHC clinical settings. We were also able to determine our capability to estimate the cross-sectional seroprevalence within Texas’s FQHC clinical settings. The crude positivity prevalence for SARS-CoV-2 antibodies in this sample was 26.1% indicating potentially high exposure to COVID-19 for FQHC clinic employees and patients. We also demonstrated feasibility and capability to determine the presence of IgG antibodies to SARS-CoV-2 in populations with and without previous COVID-19 positive diagnosis. The inclusion of COVID-19 positive and negative participants is important as it has been a limitation of other studies and allows us to more accurately determine the seroprevalence and human response over time in a diverse representative population. Therefore, ability to determine antibodies in individual with no previous history of COVID-19 over time is a unique aspect of our program approach that may inform understanding of the timing of neutralizing antibodies across a 6-month period; current estimate indicate antibodies may be stable for 5-7 months after SARS-CoV-2 infection^13^.

Although self-reported the COVID-19 test positivity and self-report of symptoms allows us to better determine the cycle and decline of antibody levels in a large sample of Texans over a 6-month period. It is estimated that over one-third of patients that have recovered from COVID-19 have antibodies given mild or asymptomatic disease,^11^ it is important to note that in our sample, 68.7% of those with a previous positive COVID-19 test had a positive SARS-CoV-2 antibody test.

The timing of the data collection from the start of the first reported cases in Texas was approximately 6 months from the start of our data collection. The positive cases will be monitored for decline of antibody levels and collection of additional COVID-19 testing, positive results and symptoms over a further six-month enrollment period. Although the highest neutralizing antibody titers are found in severe disease,^19^ the expected waning of antibody presence is yet unknown. We posit that the presence of antibodies will vary by populations, previous exposures and symptoms. The design of our program allows us to collect survey data to best identify the demographic and clinical characteristics associated with seroprevalence response across a large state. It is estimated that there may be 10 times more SARS-CoV-2 infections than the number of reported cases^14^. Understanding the presence of antibodies in a large sample of diverse populations with and without COVID-19 diagnosis may also be used to inform state-wide initiatives, vaccination distribution and restrictions across large populations.

The Phase I setting is important to consider as we enrolled FQHCs to participate in the program to determine the presence of antibodies in both employees and patient populations. Given the predicted long-term health consequences of COVID-19 ^19^ the Texas CARES program focuses on reach of populations that are underinsured and likely to have co-morbid chronic conditions. The inclusion of this population will allow for the identification of percentage of high-risk patients with antibodies, informing their long-term care for cardiovascular, pulmonary, neurologic and emotional well-being. These data will allow for informed planning by FQHCs and state leaders to determine and address vulnerable patient population needs and for the development of interventions and strategies to best care to mitigate poor health effects of COVID-19 over time.

Among this sample, we found that our adjustments indicate that male patients may have a higher proportion of positivity for antibodies, likely due to greater exposure to COVID-19 by industry and continuation of work during restriction periods. This finding aligns with positive proportions of COVID-19 found in males as well as lower antibody levels found in women.^31^ Additionally, it is important to note the successful reach and high survey completion rate as a result of our engagement and communication strategies designed using a participatory approach to support community-academic partnerships. The engagement of FQHCs who primarily serve vulnerable populations disproportionately impacted by COVID-19 was purposeful as it allowed for reach and determination of antibody response in highly vulnerable patients.

This report has several limitations. First, the participants are voluntary and are not a representative sample of Texas residents. However, the sample represents patients and populations in three counties and areas with varied COVID-19 infection rates. Second, the data collection for COVID-19 test positivity are self-reported, however, we believe the pandemic and impact on communities increases reliability of self-reported testing and positive diagnosis. As the TX CARES sample increases we hypothesize the prevalence of antibody positive will decrease as the Phase I population represented three specific FQHC clinic settings and communities. Third, this sample was primarily women, representing the employee demographic of FQHCs and patient populations within the clinics.

Nevertheless, these findings suggest the feasibility to recruit participants from high-risk populations seeking care at FQHCs and employees serving the population. We also found that the high proportions of survey completion point to interest in the population to engage in research to identify antibody status.

This program was designed to identify the humoral immune response to SARS-CoV-2 infection in a large sample over time and may assist in determining potential vulnerability to a surge in COVID-19 cases across a large state population. We found a high estimation of seroprevalence in this first phase of our program using a high specificity and sensitivity assay in a primarily White Hispanic population. Estimating seroprevalence is important given the potential for reinfection and severity of COVID-19 in vulnerable populations with co-morbidities while vaccination uptake and reach across a state continues.

## Data Availability

Data dashboard is available at our survey website:https://sph.uth.edu/projects/texascares/

## Summary Box

In 1 or 2 sentences for each, answer the following: What is already known on this topic? What is added by this report? and What are the implications for public health practice? These answers contain the key public health message, as well as the justification for the publication. Total word limit should be no more than 75‒100 words. Answers longer than 100 words will be edited to meet the word limit. PCD does not require a summary box for every article type

## What is already known on this topic?

Infection rates of SARS CoV-2 are documented across the world, however, estimates of true infection and “natural” immunity are still unclear.

## What is added by this report?

This survey allows us to better understand “natural” immunity and exposure in a underserved population receiving care at Federally Qualified Health Centers (FQHCs) across the state of Texas. TX CARES also contributes to our understanding of engagement of underserved communities using strategies such as champions at the FQHC sites.

## What are the implications for public health practice?

Implications of this work include greater understanding of seroprevalence response as well as exposure across ages 5-80 years at FQHCs. Estimating seroprevalence is important for public health practices given the potential for reinfection and severity of COVID-19 in vulnerable populations with co-morbidities while vaccination of a larger portion of the population continues.

## Acknowledgements

This work was supported by the Texas Department of State Health Services and the University of Texas System. This analysis would not have been possible without the partnership of many. The TX CARES investigation team would like to thank our federally qualified health center partners and the Texas Association for Community Health Centersfor assisting with sharing information with families about this survey.

PCD follows ICMJE style for references

## References

1. Centers for Disease Control and Prevention. National notifiable disease surveillance system (NNDSS). Centers for Disease Control and Prevention Web site. https://wwwn.cdc.gov/nndss/conditions/coronavirus-disease-2019-covid-19/. Accessed October 10, 2020.

2. Mizumoto K, Kagaya K, Zarebski A, Chowell G. Estimating the asymptomatic proportion of coronavirus disease 2019 (COVID-19) cases on board the Diamond Princess cruise ship, Yokohama, Japan, 2020. Euro Surveill. 2020;25(10). doi:10.2807/1560-7917.ES.2020.25.10.2000180

3. Kimball A, Hatfield KM, Arons M, et al; Public Health—Seattle & King County; CDC COVID-19 Investigation Team. Asymptomatic and presymptomatic SARS-CoV-2 infections in residents of a long-term care skilled nursing facility—King County, Washington, March 2020. MMWR Morb Mortal Wkly Rep. 2020;69(13):377–381. doi:10.15585/mmwr.mm6913e1

4. Rosenberg ES, Tesoriero JM, Rosenthal EM, et al. Cumulative incidence and diagnosis of SARS-CoV-2 infection in New York. Ann Epidemiol. 2020;48:23-29.e4. doi:10.1016/j.annepidem.2020.06.004

5. Lu X, Zhang L, Du H, Zhang J, Li YY, Qu J, et al. SARS-CoV-2 Infection in Children. N Engl J Med. 2020;382(17):1663–5.

7. Ludvigsson JF. Systematic review of COVID-19 in children shows milder cases and a better prognosis than adults. Acta Paediatr.109(6):1088–95.

9. Feldstein LR, Rose EB, Horwitz SM, et al. Multisystem Inflammatory Syndrome in U.S. Children and Adolescents. N Engl J Med. 2020 Jul 23;383(4):334–346. doi: 10.1056/NEJMoa2021680. Epub 2020 Jun 29.

10. Rowley AH. Understanding SARS-CoV-2-related multisystem inflammatory syndrome in children. Nat Rev Immunol. 2020;20(8):453–454. doi:10.1038/s41577-020-0367-5.

11. Havers FP, Reed C, Lim T, et al. Seroprevalence of antibodies to SARS-CoV-2 in 10 sites in the United States, March 23-May 12, 2020. JAMA Intern Med. 2020.

12. Sutton D, Fuchs K, D’Alton M, Goffman D. Universal screening for SARS-CoV-2 in women admitted for delivery. N Engl J Med. 2020;382(22):2163–2164.

13. Ripperger TJ, Uhrlaub JL, Watanabe M, et al. Orthogonal SARS-CoV-2 serological assays enable surveillance of low-prevalence communities and reveal durable humoral immunity. Immunity. 2020;53(5):925-933.e4.

14. Stephens DS, McElrath MJ. COVID-19 and the path to immunity. JAMA. 2020;324(13):1279–1281.

15. Mahajan UV, Larkins-Pettigrew M. Racial demographics and COVID-19 confirmed cases and deaths: A correlational analysis of 2886 US counties. J Public Health (Oxf). 2020;42(3):445–447.

16. Nguyen LH, Drew DA, Graham MS, et al. Risk of COVID-19 among front-line health-care workers and the general community: A prospective cohort study. Lancet Public Health. 2020;5(9):e475–e483.

17. Moscola J, Sembajwe G, Jarrett M, et al. Prevalence of SARS-CoV-2 antibodies in health care personnel in the new york city area. JAMA. 2020;324(9):893–895.

18. Hu B, Guo H, Zhou P, Shi ZL. Characteristics of SARS-CoV-2 and COVID-19. Nat Rev Microbiol. 2021 Mar;19(3):141–154. doi: 10.1038/s41579-020-00459-7. Epub 2020 Oct 6.PMID: 33024307

19. Robbiani DF, Gaebler C, Muecksch F, et al. Convergent antibody responses to SARS-CoV-2 in convalescent individuals. Nature. 2020;584(7821):437–442.

20. Del Rio C, Collins LF, Malani P. Long-term health consequences of COVID-19. JAMA. 2020.

21. The Commonwealth Fund. David C. Radley, Sara R. Collins, and Jesse C. Baumgartner https://2020scorecard.commonwealthfund.org/state/texas/. Accessed January 20, 2021.

22. Pineles BL, et al. Racial-ethnic disparities and pregnancy outcomes in SARS-CoV-2 infection in a universally-tested cohort in Houston, Texas. Eur J Obstet Gynecol Reprod Biol. 2020. PMID: 32950276

23. Johns Hopkins University & Medicine. Coronavirus Resource Center. https://coronavirus.jhu.edu/us-map. Accessed February 23, 2021.

24. Texas Association of Community Health Centers. Programs and Services. https://www.tachc.org/Online/About/Online/About/. Accessed October, 3, 2021.

25. PhenX Toolkit. COVID-19 Protocol Library. https://www.phenxtoolkit.org/covid19. Accessed September 9, 2020.

26. Cobas. Elecsys® Anti-SARS-CoV-2. https://diagnostics.roche.com/us/en/products/params/elecsys-anti-sars-cov-2.html Accessed July 22, 2020.

27. The American Association for Public Opinion Research. 2016. Standard Definitions: Final Dispositions of Case Codes and Outcome Rates for Surveys. 9th edition. AAPOR

28. Centers for Disease Control and Prevention, National Center for Chronic Disease Prevention and Health Promotion. Behavioral Risk Factor Surveillance System. https://www.healthypeople.gov/2020/data-source/behavioral-risk-factor-surveillance-system Accessed December 26, 2020.

29. Frankel, JA, Froot, KA. Using Survey Data to Test Standard Propositions Regarding Exchange Rate Expectations. The American Economic Review. Vol. 77, No. 1 (Mar., 1987), pp. 133–153 (21 pages)

30. Kaplowitz MD, Hadlock TD, Levine R. A comparison of web and mail survey response rates. Public Opinion Q. 2004;68(1):94–10

31. Gudbjartsson DF, Helgason A, Jonsson H, et al. Spread of SARS-CoV-2 in the Icelandic population. N Engl J Med. 2020;382(24):2302–2315.

